# Bulwark Effect of Response in a Causal Model of Disruptive Clinician Behavior

**DOI:** 10.1101/2024.06.03.24308343

**Authors:** Manabu Fujimoto, Mika Shimamura, Hiroaki Miyazaki, Kazuto Inaba

## Abstract

**INTRODUCTION:** Disruptive clinician behavior (DCB) refers to unethical and unprofessional behavior that seriously affects patient safety by disrupting relationships among healthcare professionals and causing dysfunctional communication and teamwork. DCB often persists as an organizational culture in Japanese healthcare settings because of problems in the conventional leadership system along with professional and positional hierarchies. Therefore, this study verified a causal model of DCB in Japanese healthcare, including triggers, response, and impact.

**METHODS:** Staff at two general hospitals (751 and 661 beds) were surveyed using a web-based questionnaire. In total, 256 staff who had experienced victimization and agreed to complete the questionnaire were included in this study. The questionnaire comprised demographic information, a DCB scale, and items covering causal indicators of DCB: triggers, response, and impact (psychological/social and medical/management).

**RESULTS:** Mediation and moderated mediation analyses showed that: (1) DCB had a negative impact on the medical/managerial state, which was partially mediated by psychological/social impact; and (2) the responses of victims and others acted as a bulwark in reducing the psychological/social impact to some extent.

**DISCUSSION:** A prompt response to DCB as a bulwark reduces victims’ psychological and social adaptation deterioration. Therefore, occurrences of DCB should not be overlooked, and the victim and those around them should respond positively. However, response as a bulwark cannot protect the organization’s medical care and management. Therefore, it is important to prevent DCB.

---

Disruptive clinician behavior (DCB) refers to unprofessional and unethical behavior that seriously affects patient safety by disrupting interpersonal relationships among healthcare providers and resulting in communication and teamwork dysfunction.^1,2^ Disruptive interpersonal behaviors among healthcare providers harm communication and teamwork among providers, but the types of behaviors are diverse. A breakdown of disruptive behaviors showed physical violence accounted for less than 5%, but there were high incidences of abusive language, verbal abuse, rudeness, scolding, abusive anger, insults, and threats.^3^ Other categories of such behaviors include sexual harassment, racism, threatening behavior, neglect, disrespect, throwing of items, inappropriate documentation, and refusal to attend meetings.^4,5^ The Disruptive Behavior Questionnaire (PASCAL METRICS, Inc.) can be used to assess different categories of disruptive behaviors, including: divisive behavior, intimidating behavior, disrespectful behavior, inhibiting behavior, and offensive behavior. Petrovic et al. identified the top 20 disruptive behaviors in a literature review, which they classified into eight categories: passive-aggressive behavior, physical or verbal threats, verbal abuse, physical violence, harassment, intimidation, bullying, and discrimination.^6^ However, these categories have not been statistically substantiated. Fujimoto et al. developed a psychological scale based on free descriptions from witnesses and victims of DCB according to the scale construction procedure and confirmed its validity and reliability.^7^ Those authors showed that DCB has a hierarchy structure (Table 1), and may be divided into interpersonal and job-related aggression. The former category, interpersonal aggression, consists of psychological aggression, incivility, ignoring, and physical violence. Psychological aggression was further subdivided into intimidation, reproof, threats, and abusive language. Job-related aggression comprises mismanagement practices and passive aggression. Mismanagement practices were further divided into non-supportive coercion and arbitrary decisions. As discussed above, DCB can take various forms in interpersonal behavior and performance of duties.

**Table 1.**
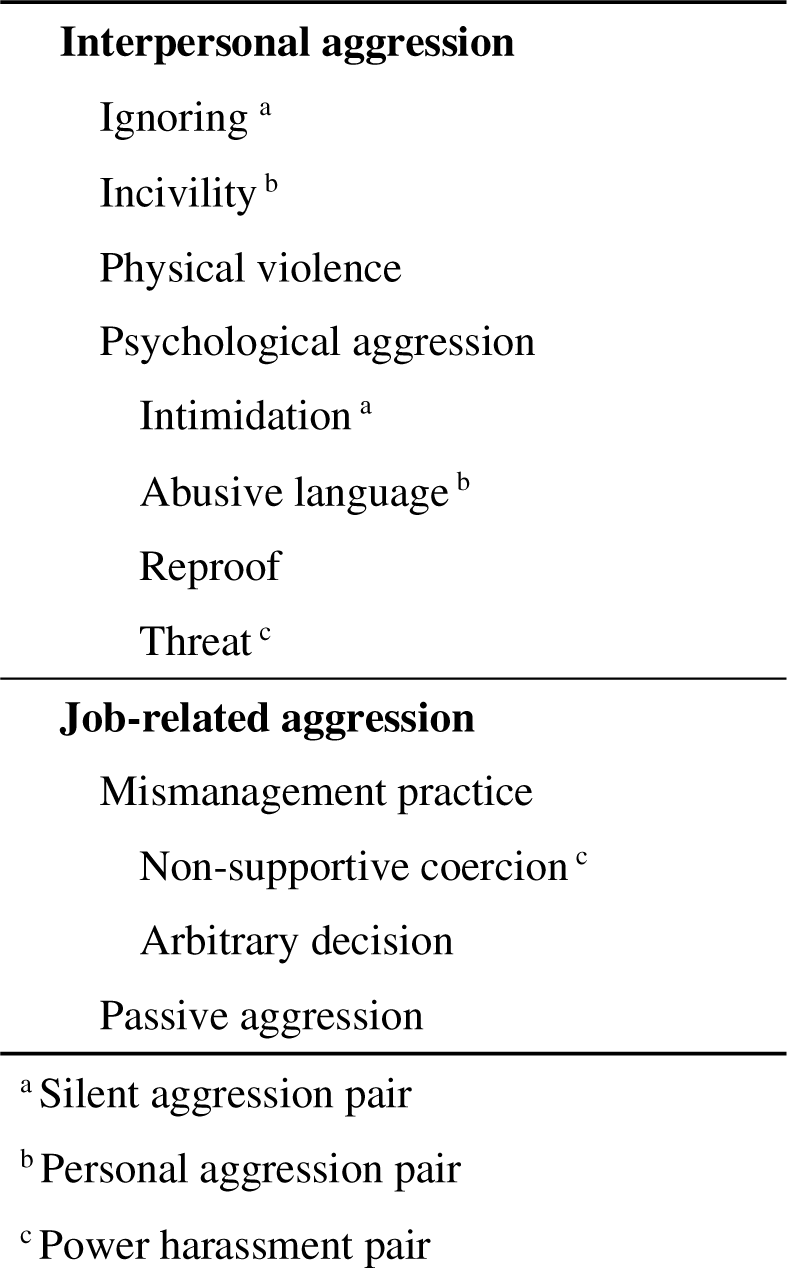
Empirical categories of disruptive clinician behavior.

|  |  |
| --- | --- |
| **Interpersonal aggression** |  |
|  | Ignoring <sup>a</sup> |
|  | Incivility <sup>b</sup> |
|  | Physical violence |
|  | Psychological aggression |
|  | Intimidation <sup>a</sup> |
|  | Abusive language <sup>b</sup> |
|  | Reproof |
|  | Threat <sup>c</sup> |
| **Job-related aggression** |  |
|  | Mismanagement practice |
|  | Non-supportive coercion <sup>c</sup> |
|  | Arbitrary decision |
|  | Passive aggression |
| <sup>a</sup> Silent aggression pair |  |
| <sup>b</sup> Personal aggression pair |  |
| <sup>c</sup> Power harassment pair |  |

DCB occurs routinely in healthcare settings.^8^ For example, 71% of hospital administrators reported that DCB occurred at least monthly, and 11% said this occurred daily. Since 2000, DCB has been viewed as a problem in the US; the Joint Commission adopted a zero-tolerance policy and is actively working to eradicate DCB.^9^ In contrast, awareness of DCB is low in Japan, and many people believe it is inevitable that they will be subjected to DCB.^10^ In this context, problems with the causal leadership system and hierarchy have been highlighted. The same is true in the US, where not all DCB is perpetrated with clear intent to harm. There is a small number of healthcare providers who unknowingly engage in violent and often angry behavior that affects interpersonal relationships and the quality of healthcare. ^8,11,12^ One of the few empirical studies focused on DCB in Japan found that this behavior often included explicit and spiteful behavior.^13^ Explicit DCB may be performed to vent anger or frustration. As such, it is expressed as physical violence or psychological aggression. Concerningly, organizations tend to overlook physicians at the top of the professional hierarchy, even when they engage in explicit DCB. ^8,14^ Spiteful DCB is tantamount to bullying in the workplace and often takes the form of incivility and ignoring. Many previous studies indicated that DCB among nurses, who make up the majority of healthcare workers, generally takes the form of bullying.^15,16,17^

This study focused on Japanese healthcare providers. Well-known Japanese cultural characteristics include group orientation, sociability, and wholeness.^18^ Furthermore, historically, Confucian morality is solid and hierarchical relationships are strict.^19^ These characteristics mean that peer pressure and synchronized behavior are likely to occur, and members are unlikely to disagree with an organization’s norms and policies; if they rebel, they are immediately subjected to social sanctions, such as being ostracized. Because of these cultural characteristics, Japanese organizations tend to be constitutionally psychologically insecure, which means that their members do not freely express opinions and ideas without fear of blame or criticism. This concept has recently been emphasized in the medical world,^20^ as organizations with higher levels of psychological safety show higher performance than those with lower levels of psychological safety.^21^ Given that team medicine has become mainstream, it is assumed that if medical professionals are guaranteed an environment in which they can freely express their opinions, DCB will decrease and the quality and safety of medical care will improve. It has been noted that Japanese society is feudalistic; however, other countries are similar, especially in medical institutions.^22^ The shared cause of saving patients’ lives fosters an atmosphere in which anything is acceptable. A study focused on Japanese medical professionals is expected to provide essential insights in elucidating the reality of DCB that occurs in medical organizations that tend to be closed.

Based on the above research background, this study attempted to elucidate a causal model of DCB in the Japanese medical field. The Johns Hopkins model for DCB has four serially connected components: triggers, DCB, response, and impact (Figure 1). Demonstrating the relevance of these components was one of the main tasks of our study. Regarding the model components, the impact of DCB has adverse effects on both the organization^23,24^ and the victim.^25,26^ Therefore, we treated impact discriminately. In addition, knowing how to respond when DCB occurs is essential.^27,28^ For example, Rosenstein recommended a proactive approach to early detection and intervention and improving organizational culture because of the complexity of the nature, causes, and effects of disruptive behavior.^4^ Therefore, this study focused on the effects of response to DCB in validating the model.

**Figure 1.**
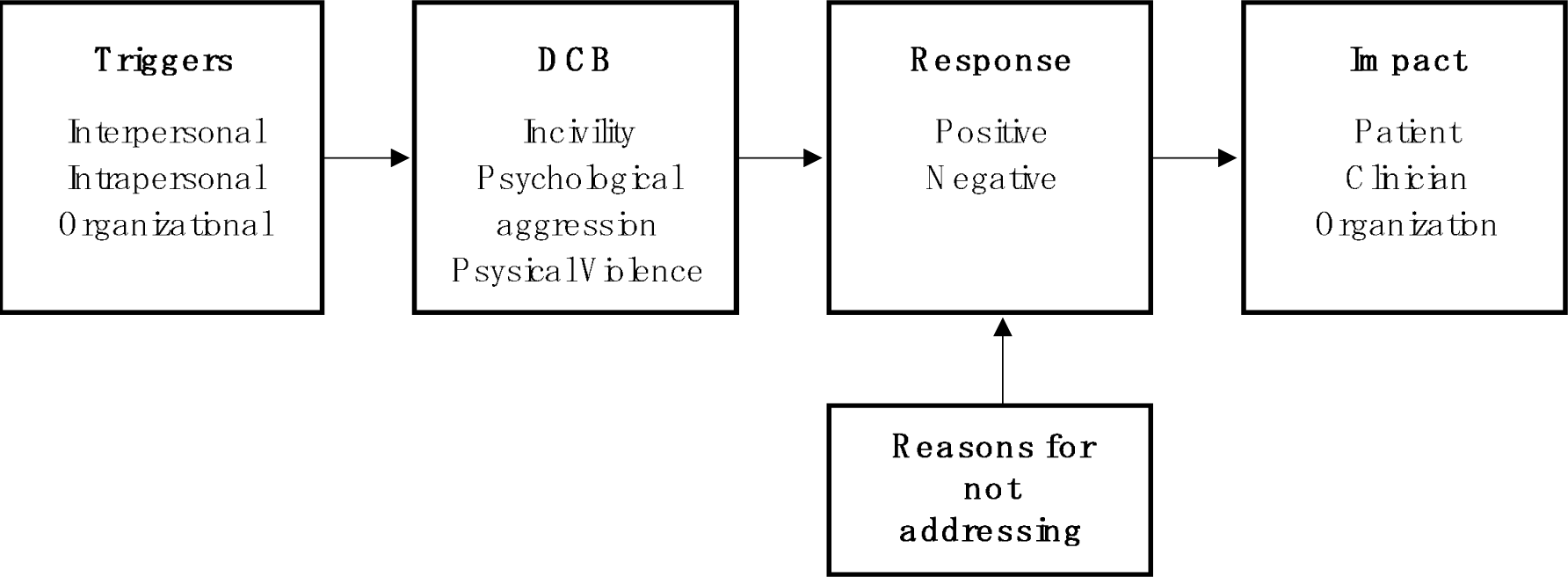
Johns Hopkins model for disruptive clinician behavior Copyright© 2012, The Johns Hopkins Health System Corporation. All rights reserved.

There are existing scales for measuring DCB.^7^ In this study, we collected information about triggers, response, and impact using items in an open-ended questionnaire that was organized and categorized. Our analysis tested a hypothesized model of causality for DCB. We employed an adjusted mediation model to clarify the response effect for the analytical model. The difference between this analytical model and the Johns Hopkins model is that the statistical analysis treated “response” as an adaptation variable. First, a direct path was set for the triggers-DCB-negative impact on the organization, with reference to the Johns Hopkins DCB model. Next, consistent with the definition of DCB given in the introduction, we added an indirect path to the DCB-negative impact on the organization that mediated the negative impact on the victim because DCB has a serious impact on patient safety by disrupting healthcare providers’ relationships and making communication and teamwork dysfunctional. Finally, to apply the response component in the Johns Hopkins model to our analytical model, arrows representing the response were drawn on the paths from DCB to the two impacts and between the two impacts.

## METHODS

### Preliminary Survey

An open-ended Internet-based questionnaire was administered to 712 nursing professionals nationwide regarding their experiences of witnessing or being victimized by DCB. The questionnaire items covered DCB, triggers, response, and impact. The responses regarding behavior were drawn from the “Psychological Scale for Measuring DCB”.^7^ The descriptions collected for triggers, responses, and impacts were first summarized and then categorized using content analysis. Next, the responses were finalized after modification by the co-authors, who had significant experience in medical safety. In this process, triggers were categorized into six indicators and 12 items, and responses were categorized into eight indicators and 16 items. Impact was dichotomized as a negative impact on either the organization or the victim. The former referred to the medical/ managerial impact, which reflected deterioration of the quality of medical care provided by the hospital and the hospital management. The latter comprised psychological/social impact, which referred to deterioration of the psychological and social adaptation of the victims. Each of these impacts was classified using three indicators and six items.

### Procedures and Participants

A field survey using the Google Forms platform was administered to the staff of two general hospitals with 751 and 661 beds, respectively. Of the 446 participants who agreed with the purpose 6 had experienced victimization by DC

### Survey Items

First, participants were asked to respond to five items covering demographic information. Next, they completed the 38-item Psychological Scale for Measuring DCB,^7^ which comprises 10 indicators. Participants were asked to rate the degree to which they had experienced each DCB in the case using a 6-point scale (from “not at all” to “always”). Finally, participants completed the Inventory on Causal Variable of DCB, which comprehensively examined triggers, response, psychological/social impact, and medical/managerial impact. Responses were on a six-point scale (from “not at all” to “very much”) to determine the degree of applicability.

### Statistical Analyses

Multivariate analysis of variance was used to examine the differences between the data for the two hospitals. First, confirmatory factor analysis and calculation of the omega coefficient were used to check the reliability of the item sheets for the DCB causal variables. Next, principal component analysis was used to compress the factors for each indicator. Structural equation modeling (SEM) was then used to validate the direct path model. Furthermore, mediation analysis was used to examine the psychological/social impact of the mediation effects. Finally, we tested the adaptation effect of response through adjusted mediation analysis.

### Ethical Considerations

This study adhered to the relevant research ethics guidelines after obtaining approval from Human Research Ethics Committee of Ritsumeikan University (衣笠-人-2022-106) and Research Ethics Committee of JCHO Chukyo Hospital (あああああ). After obtaining approval from the hospital administrators, a written request for research cooperation was distributed to hospital staff. This document clearly stated the purpose and methods of the study, clarified that participation was voluntary, and noted data would be statistically processed so that individuals could not be identified. Staff who agreed to cooperate in this study accessed the Google Forms questionnaire using the URL provided in the request document.

## RESULTS

### Confirmation of Differences Between Hospitals

To confirm the differences between data for the two hospitals, we performed multivariate analysis of variance with the hospital as the independent variable and scale data as the dependent variables. The multivariate test was non-significant (Wilks’s L=0.847, F(30, 225)=1.358, p=.111, partial h2=.153). Therefore, the data from the two hospitals were combined for the subsequent analyses.

### Confirming the Reliability of the Indicators

We tested the reliability of the 20 indicators for the DCB causal variables in the item form. Each indicator had two items, and their respective omega coefficients ranged from .651 to .914, except that for avoidance, which was low at .480 (Table 3). Confirmatory factor analysis was conducted after the scale scores were calculated. The results showed that triggers (goodness of fit index [GFI]=.965, comparative fit index [CFI]=.961, root mean square error of approximation [RMSEA]=.095), impact (GFI=.953, CFI=.979, RMSEA=.092), and response (GFI=.933, CFI=.962, RMSEA=.073) all showed a certain degree of fit. In terms of the impact, we used a model in which correlation paths were drawn between the observed variables representing the psychological/social impact and medical/managerial impact factors.

### Compression of Indicators

We used principal component analysis to compress the indicators in the mediation and first principal component, except for physical violence, which had a significantly low mean value. For response, five indicators were highly loaded on the first principal component, except for three negative factors (avoidance, servile submission, and adulation). Therefore, “response” in the analytical model meant a positive response. All indicators for triggers, psychological/social impact, and medical/managerial impact were highly loaded on the first principal component. We therefore used the first principal component scores for all variables in the subsequent analyses.

### Validation of DCB Triggers and Impact

SEM was used to test the causal model of triggers-DCB-medical/managerial impact. The direct path from reasons to destructive behavior and DCB to medical/managerial impact were significant, with estimates of .619 (standard error [SE]=.049, p<.001) and .556 (SE=.052, p<.001), respectively.

### Examining the Indirect Effects of Psychological/Social Impact

We used mediation analysis to test the model, including indirect paths (Figure 2). The results showed that the direct path from DCB to medical/managerial impact had an estimated value of .464 (SE=.058, p<.001), the indirect path from DCB to psychological/social impact had an estimated value of .486 (SE=.055, p<.001), and the indirect path from psychological/social impact to medical/managerial impact had an estimated value of .190 (SE=.058, p=.001). To verify the mediating effect, we calculated 95% confidence intervals using the bootstrap method (sample size: 2000). The results were significant, with an estimate of .092 (SE=.032, p=.001; lower bound=.043, upper bound=.148). The partial mediation model was supported as the direct path estimate from DCB to medical/managerial impact was somewhat reduced from .556 to .464 by the mediation of psychological/social impact.

**Figure 2.**
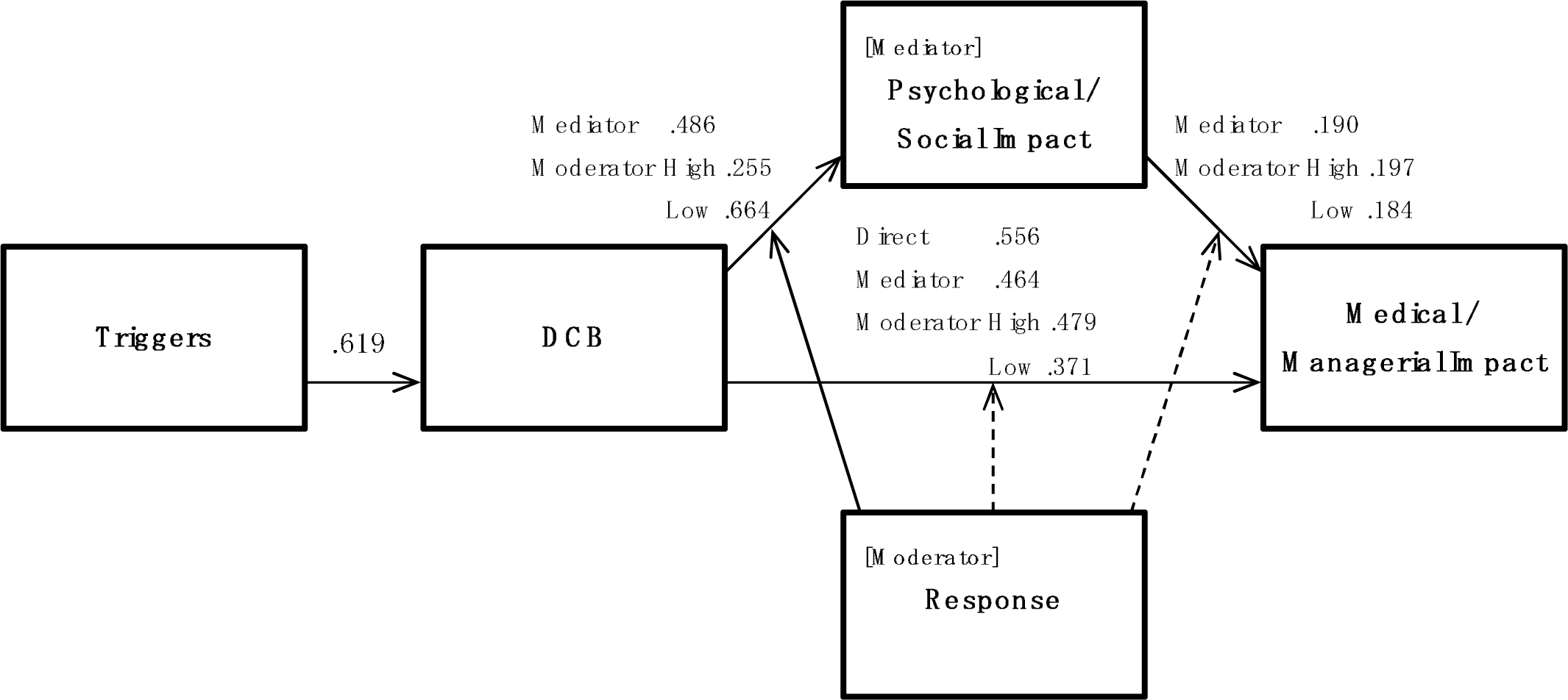
Causal model of DCB and estimated value of each path. Causal model of disruptive clinician behavior and estimated value of each path. Mediator values are estimates of paths obtained by mediation analysis and moderator values are estimates of paths obtained by an adjusted mediation analysis. Dotted lines indicate little adaptation effect.

### Verification of Moderate Effects of Response

We tested the moderating effect of response through a moderated mediation analysis (Figure 2). First, we considered the paths for response. The path from response to medical/managerial impact was estimated at .125 (SE=.055, p=.025) and that from response to psychological/social impact was estimated at .133 (SE=.057, p=.020). The path from the interaction term between DCB and response to medical/managerial impact was estimated at .017 (SE=.046, p=.746), that from the interaction term between DCB and response to psychological/social impact was estimated at −.204 (SE=.047, p<.001), and that from the interaction term between psychological/social impact and response to medical/managerial impact was estimated at .006 (SE=.051, p=.908).

Next, for the paths in the mediation models for DCB, medical/managerial impact, and psychological/social impact, the direct path from DCB to medical/managerial impact was .479 for the high response group (+1 standard deviation [SD]). The direct path from DCB to medical/managerial impact was .479 (SE=.071, p<.001). The indirect path from DCB to psychological/social impact was .255 (SE=.072, p<.001), the indirect path from psychological/social impact to medical/managerial impact was .479 (SE=.071, p<.001), and the indirect path from psychological/social impact to medical/managerial impact was .197 (SE=.078, p=.012). Conversely, for the low response group (−1 SD), the direct path from DCB to medical/managerial impact was .371 (SE=.087, p<.001), the indirect path from DCB to psychological/social impact was .664 (SE=.076), and the indirect path from psychological/social impact to medical/managerial impact was .184 (SE=.080, p=.023).

Finally, we tested the indirect effects using the bootstrap method. The results for the high response group (+1 SD) showed an estimated value of .055 (SE=.026, p=.034, 95% lower bound=.015, 95% upper bound=.121) and those for the low response group (−1 SD) showed an estimated value of .118 (SE=.053, p=.027, 95% lower bound=.016, 95% upper bound=.226), and were significant. Estimates for the direct path from DCB to medical/managerial impact were significantly lower in the low group (.494–.371) compared with the high group (.528–.479). Both the high and low groups supported the partial mediation model. However, the mediation effect was superior in the low group compared with the high group.

## DISCUSSION

This study confirmed that personality and competence issues of perpetrators and victims and workplace climate issues can cause destructive behavior among medical personnel (triggers-DCB). Furthermore, we confirmed that DCB directly harmed the quality of healthcare and hospital management and indirectly endangered healthcare and management by worsening the victim’s psychological and social adaptation (mediating effect of psychological/social impact). We also found that when DCB occurred, it was effective for the victim and those around them to respond appropriately to protect the victim’s psychological and social adaptation (bulwark effect: moderating effect of response). In particular, our results suggested that it was effective when the victim sought social support from those around them and when those around them started to solve the problem. However, response as a bulwark only blocked the indirect path to psychological/social impact. Therefore, the effects of DCB flowed back to the direct path, which more directly damaged patient safety and hospital management. In other words, the responses of the victims and those around them did not prevent adverse effects on medical and managerial conditions. In summary, there were two main findings from this study. The first finding was that it is crucial not to overlook DCB and ensure that responses to any incidents are appropriate to create an environment where medical personnel can continue to work with good well-being. However, regardless of the response, DCB damages the quality of hospital care and management. The second main finding was the need to prevent the occurrence of DCB to protect the quality of healthcare and management of the organization. Based on these findings, the following discussion focused on responding to and preventing DCB.

Interpersonal DCB can be dichotomized into overt and spiteful forms, but overt DCB, such as violence and verbal abuse, directly reduce the quality of healthcare.^13^ These often occur as administrative sanctions in hierarchical relationships, educational reprimands in teaching relationships, or abuse from physicians to nursing staff/co-medicals as part of performing their duties as higher-ranking personnel.^29^ Personal circumstances, such as the perpetrator’s personality, mood, or physical condition at the time, may also be triggers.^30^ Therefore, it is necessary to raise awareness so people understand that DCB is an “old idea” that needs to be updated. Updating awareness is not limited to the perpetrators; the organizational culture must be improved as it is a problem for the entire organization to prevent DCB from being overlooked as “something that cannot be helped” if it is observed.

Explicit DCB tends to be more apparent, and violence and sexual crimes tend to be less common, as they may violate criminal law.^13^ However, spiteful DCB, such as bullying and neglect, tend to be overlooked.^31^ Spiteful DCB worsens victims’ psychological and social adaptation status.^32^ As a result, victims have difficulty performing their regular duties, which affects patient outcomes^33^ or results in a decision to leave their jobs.^34^ Furthermore, the quality of medical care and the management of the medical institution suffer significant damage.^35^ Employees are valuable assets for healthcare organizations, especially as staffing shortages are severe in healthcare institutions worldwide.^36^ As demonstrated in this study, protecting victims from the consequences of DCB by appropriate responses can reduce deterioration of their psychological and social adaptation status. As identified in the preliminary study (Table 2), specific responses are for the victim to cope directly or request instrumental and emotional support from those around them, who can act toward a solution or mediate between the victim and perpetrator.

**Table 2.** Participants’ demographic data.

|  | Hospital A | Hospital B |
| --- | --- | --- |
| Physician | 20 | 11 |
| Nursing professional | 39 | 101 |
| Paramedic | 21 | 39 |
| Non-medical professional | 5 | 22 |
| Female | 49 | 123 |
| Male | 35 | 45 |
| Unanswered | 1 | 5 |
| Age (years) 20s | 14 | 21 |
| 30s | 21 | 41 |
| 40s | 26 | 51 |
| 50s | 17 | 36 |
| 60s | 5 | 16 |
| Unanswered | 2 | 8 |
| Total | 85 | 173 |

**Table 3.** Results of principal component analysis of disruptive clinician behavior, triggers, response, and impact.

|  |  |  | *M* | *SD* | *w* |
| --- | --- | --- | --- | --- | --- |
| **DCB** | 1st principal component contribution rate | 35.03% |  |  | .933 |
| **Ignoring** | 1st principal component loading | .605 | 2.51 | 1.50 |  |
| **Incivility** |  | .751 | 3.25 | 1.57 |  |
| **Reproof** |  | .839 | 3.03 | 1.50 |  |
| **Threat** |  | .837 | 3.79 | 1.50 |  |
| **Intimidation** |  | .739 | 2.67 | 1.44 |  |
| **Abusive language** |  | .845 | 3.50 | 1.54 |  |
| **Physical violence** |  | .351 | 1.24 | 0.69 |  |
| **Non-supportive coercion** |  | .684 | 1.77 | 1.02 |  |
| **Arbitrary decision** |  | .810 | 2.30 | 1.22 |  |
| **Passive aggression** |  | .746 | 2.41 | 1.31 |  |
| **Triggers** | 1st principal component contribution rate | 35.03% |  |  | .789 |
| **Perpetrator competence** | 1st principal component loading | .575 | 2.80 | 1.47 | .753 |
|  | The perpetrator lacked knowledge or experience in the job |  | 2.44 | 1.45 |  |
|  | The perpetrator lacked competence and aptitude for the job |  | 3.17 | 1.80 |  |
| **Perpetrator's personality** | 1st principal component loading | .669 | 3.84 | 1.43 | .651 |
|  | The perpetrator had communication and socialization problems |  | 4.28 | 1.59 |  |
|  | The perpetrator had a lax and inconsiderate personality |  | 3.40 | 1.74 |  |
| **Victim's competence** | 1st principal component loading | .477 | 2.64 | 1.29 | .859 |
|  | The victim lacked knowledge or experience in the job |  | 2.75 | 1.44 |  |
|  | The victim lacked competence or aptitude for the job |  | 2.53 | 1.32 |  |
| **Victim's personality** | 1st principal component loading | .556 | 1.93 | 0.99 | .782 |
|  | The victim had problems with communication and socializing |  | 2.16 | 1.21 |  |
|  | The victim had a lax and inconsiderate personality |  | 1.70 | 0.99 |  |
| **Press of business** | 1st principal component loading | .518 | 3.42 | 1.76 | .878 |
|  | There was a regular shortage of staff at the workplace |  | 3.46 | 1.89 |  |
|  | Busy or urgent situation |  | 3.38 | 1.84 |  |
| **Normalization** | 1st principal component loading | .721 | 4.02 | 1.53 | .681 |
|  | There was no one around to blame |  | 4.45 | 1.65 |  |
|  | Disruptive behavior was the norm in the workplace |  | 3.59 | 1.85 |  |
| **Response** | 1st principal component contribution rate | 34.91% |  |  | .868 |
| **Direct action by the victim** | 1st principal component loading | .488 | 2.90 | 1.52 | .854 |
| **Requested emotional support** | 1st principal component loading | .764 | 3.73 | 1.53 | .784 |
| The victim talked to others |  |  | 3.82 | 1.70 |  |
| The victim got emotional support from others |  |  | 3.64 | 1.68 |  |
| **Avoidance** | 1st principal component loading | .156 | 3.90 | 1.42 | .480 |
| The victim distanced themselves from the perpetrator |  |  | 4.03 | 1.79 |  |
| The victim tried not to care |  |  | 3.78 | 1.72 |  |
| **Servile submission** | 1st principal component loading | -.031 | 3.26 | 1.60 | .733 |
| The victim had no choice but to give up |  |  | 3.58 | 1.83 |  |
| The victim did what the perpetrator wanted them to do |  |  | 2.94 | 1.79 |  |
| **Direct action by others** | 1st principal component loading | .797 | 2.05 | 1.29 | .890 |
| Others systematically took remedial measures |  |  | 2.05 | 1.37 |  |
| Others tried to solve the problem by confronting the perpetrator and the problem |  |  | 2.05 | 1.35 |  |
| **Arbitration** | 1st principal component loading | .791 | 1.86 | 1.19 | .812 |
| Others intervened between the perpetrator and the victim and tried to mediate |  |  | 1.92 | 1.37 |  |
| Others pacified the perpetrator |  |  | 1.81 | 1.24 |  |
| **Adulation** | 1st principal component loading | .081 | 2.28 | 1.32 | .782 |
| Others did what the perpetrator wanted them to do |  |  | 2.60 | 1.57 |  |
| Others sympathized with the perpetrator |  |  | 1.95 | 1.36 |  |
| **Psychological/social impact** | 1st principal component contribution rate 80.33% |  |  |  | .923 |
| **Psychological state** | 1st principal component loading | .919 | 4.13 | 1.52 | .801 |
| Became physically and mentally unwell |  |  | 3.99 | 1.69 |  |
| Lost motivation to work |  |  | 4.26 | 1.65 |  |
| **Quality of care** | 1st principal component loading | .852 | 3.00 | 1.61 | .789 |
| Became confused and unable to think calmly |  |  | 3.29 | 1.83 |  |
| Impaired ability to provide medical care |  |  | 2.70 | 1.73 |  |
| **Interpersonal relationships** | 1st principal component loading | .917 | 3.96 | 1.51 | .755 |
| Communication became difficult |  |  | 4.54 | 1.52 |  |
| Difficulty staying in the workplace |  |  | 3.39 | 1.84 |  |
| **Medical/managerial impact** | 1st principal component contribution rate 69.47% |  |  |  | .875 |
| **Quality of medical care** | 1st principal component loading | .892 | 3.31 | 1.62 | .768 |
| The workload increased because of staff shortages |  |  | 3.09 | 1.83 |  |
| Hindered the organization's improvement in medical quality and safety |  |  | 3.53 | 1.78 |  |
| **Hospital management** | 1st principal component loading | .785 | 2.36 | 1.34 | .749 |
| Damaged the hospital's social credibility and reputation |  |  | 2.66 | 1.68 |  |

Although the interpersonal aspect of DCB tends to be emphasized, this behavior also has a job-related aspect. Irresponsible personnel actions and unilateral decision enforcement by hospital organization managers that are unwanted by the parties involved may also be spiteful variants of DCB in that they are difficult to manifest. Poor management and guidance can reduce the victim’s work ethic.^37^ In addition, the victim may engage in passive aggression,^7^ such as skipping medical appointments, missing meetings, or refusing to perform to the best of their ability. The manager/leader will again reprimand and punish them for this explicit passive-aggressive work attitude. The victim then engages in more passive aggression. The main characteristic of job-related DCB is that the perpetrator and the victim switch places, creating a negative spiral. Breaking this chain is a critical issue for DCB prevention.

Countermeasures against DCB can also be systematic DCB against perpetrators. It is important to remember that perpetrators are as much a part of the healthcare organization as the victims. Under the zero-tolerance principle adopted in the US medical safety measures, perpetrators are punished severely to deter further problems. However, this entails similar structural problems of job-related DCB, and the system of sanctions undermines trust and cooperation within the organization.^38^ In a zero-tolerance approach, once a DCB occurs, the perpetrator becomes psychologically demotivated as their commitment to the organization and motivation for their job diminishes.

Zero-tolerance-based strict punitive measures are also organizational DCB. The more cohesive the group, the higher the performance.^39^ If the goal is for all staff members to have high well-being and work in a supportive environment, high quality and safety in healthcare will be a natural result. Therefore, the organization’s objective should be to create an environment where all medical personnel can have a tolerant mindset, build receptive relationships, and work with a high level of well-being. The study of conflict resolution is a discipline that deals with individual and social conflict issues. Rather than unilaterally restraining the perpetrator with traditional punishments (i.e., retributive justice), this discipline encourages dialogue among the perpetrator, victim, and other parties involved. It aims to rebuild interpersonal relationships, including with those involved (i.e., restorative justice).^40^ Following this lead, we should ensure a paradigm shift from retributive justice through the zero-tolerance principle to restorative justice through conflict resolution. Various interventions have been recommended to counter DCB based on a zero-tolerance principle.^27,28^ Depending on the situation, interventions for the parties involved include punishment for noncompliance, anger management, stress management, coaching, and counseling. Interventions for the organization include knowledge of DCB, awareness of policies, setting codes of conduct to create a DCB-free work environment and reporting systems for DCB, communication training for staff, and awards for measures against DCB. However, DCB continues to have severe consequences for individuals and organizations. A shift in principle toward destructive behavior among healthcare providers is now considered necessary to maximize their well-being and improve healthcare quality and hospital management.

### Challenges and Further Prospects for this Study

This study tested a causal model of DCB and confirmed the partially mediating effects of psychological/social impact and the moderating effects of a positive response. In addition, the finding that DCB directly and indirectly negatively affects the quality and management of hospital healthcare was important. However, this study was based on a psychological paradigm. Therefore, subjective evaluation measures were used in the analyses. To reinforce the findings of this study, complementary verification using objective indicators is required, such as the relationship between the number and frequency of DCB incidents occurring annually, number of users, and financial statements. Having proposed prompt responses to DCB that have occurred and organizational climate reforms to prevent DCB, we would like to conduct a further fieldwork study to formulate specific plans and verify their effectiveness in actual hospitals.

## Data Availability

All data produced in the present study are available upon reasonable request to the authors

## ACKNOWLEDGMENTS

We thank Edanz (https://jp.edanz.com/ac) for editing a draft of this manuscript.

## REFERENCES

1. Rosenstein AH. Original research: nurse-physician relationships: impact on nurse satisfaction and retention. AJN. 2002;102:26–34.

2. Martinez W, et al. Qualitative Content Analysis of Coworkers’ Safety Reports of Unprofessional Behavior by Physicians and Advanced Practice Professionals. J Patient Saf. 2021 Dec 1;17(8):e883–e889. doi:10.1097/PTS.0000000000000481.

3. Rosenstein AH. Disruptive and Unprofessional Behaviors. In: Brower KJ, Riba MB editors. Physician Mental Health and Well-Being: Research and Practice. Springer Nature; 2017. pp.61–85.doi:10.1007/978-3-319-55583-6_3

4. Rosenstein AH, Naylor B. Incidence and impact of physician and nurse disruptive behaviors in emergency department. J Emerg Med. 2012;43(1):139–148.

5. Neff K. Understanding and managing physicians with disruptive behavior. In: Ransom SB, et al., editors. Enhancing physician performance: advanced principles of medical management. Tampa (FL): American College of Physician Executives; 2000.pp.45–72.

6. Petrovic MA, Scholl AT. Why we need a single definition of disruptive behavior. Cureus. 2018 Mar 18;10(3): e2339. doi:10.7759/cureus.2339.

7. Fujimoto M, et al. Development of a Psychological Scale for Measuring Disruptive Clinician Behavior. J Patient Saf. 2023 Dec 1;19(8):564–572.

8. The Joint Commission. Behaviors that undermine a culture of safety. Sentinel Event Alert. 2008 Jul 9;(40):1–3. Available from: https://www.jointcommission.org/-/media/tjc/documents/resources/patient-safety-topics/sentinel-event/sea-40-intimidating-disruptive-behaviors-final2.pdf

9. The Joint Commission. LD_Code of Conduct Policy. 7 2020. [Online]. Available at:https://store.jcrinc.com/assets/1/7/POLB_Behavioral_Sample_Pages.pdf. Accessed August 18, 2022.

10. Nagao Y. The relationship between disruptive behavior and the quality of medical care in surgical treatment in Japan (Honpou no syujutusinryo niokeru disruptive behavior to iryonositu no kankei in Japanese).The Uehara Memorial Fundation report. 2012;28:1–6.

11. Rosenstein AH, O’Daniel M. A Survey of the Impact of Disruptive Behaviors and Communication Defects on Patient Safety. Jt Comm J Qual Patient Saf. 2008;34(8):464–471.

12. Dang D, et al. Do Clinician Disruptive Behaviors make an Unsafe Environment for patients?. J Nurs Care Qual. 2016;31(2):115–123.

13. Fujimoto M, Shimamura M, Inaba K. Influence of disruptive behaviors in Japanese medical setting. J Clin Ethics. 2021;9:29–40. Japanese.

14. MacDonald O. Disruptive Physician Behavior. American College of Physician Executives. Disruptive Physician Behavior. 2011. Available from: https://kff.org/wp-content/uploads/sites/2/2013/03/quantiamd_whitepaper_acpe_15may2011.pdf

15. Braun K, et al. Verbal abuse of nurses and non-nurses. J Nurs Manag. 1991;22(3):72–76.

16. Wilson BL, et al. Bullies at Work: The Impact of Horizontal Hostility in the Hospital Setting and Intent to Leave.J Nurs Adm. 2011;41(11):453–458.

17. Jaber HJ, et al. Perceived Relationship Between Horizontal Violence and Patient Safety Culture Among Nurses. Risk Manag Healthc Policy. 2023 Aug 14;16:1545–1553.

18. Benedict. R. The chrysanthemum and the sword: Patterns of Japanese culture. Houghton: Mifflin Company; 1946.

19. Tsuchida K. Introduction to Confucianism (Jyukyo nyumon in Japanese). Tokyo: Tokyo Daigaku Shuppankai; 2011.

20. Latessa RA, et al. Psychological safety and accountability in longitudinal integrated clerkships: a dual institution qualitative study. BMC Med Educ. 2023 Oct 12;23(1):760.

21. Edmondson AC, Lei A. Psychological Safety: The History, Renaissance, and Future of an Interpersonal Construct. Ann. Rev. Organ. Psychol. Organ. Behav.. 2014;1:23–43.

22. Gergerich E, Boland D, Scott MA. Hierarchies in interprofessional training. J Interprof Care. 2019 Sep-Oct;33(5):528-535. doi: 10.1080/13561820.2018.1538110. Epub 2018 Nov 1.

23. Rawson JV, et al. The cost of disruptive and unprofessional behaviors in health care. Acad Radiol. 2013 Sep;20(9):1074–6.

24. Lewis C. The impact of interprofessional incivility on medical performance, service and patient care: a systematic review. Future Healthc J. 2023 Mar;10(1):69–77.

25. Brewer CS, et al. Positive work environments of early-career registered nurses and the correlation with physician verbal abuse. Nurs Outlook. 2013 Nov-Dec;61(6):408–16.

26. Aunger J, et al. Drivers of unprofessional behaviour between staff in acute care hospitals: A realist review PREPRINT (Version 1). ResearchSquare. 2023.

27. Rosenstein AH. Physician disruptive behaviors: Five year progress report. World J Clin Cases. 2015 Nov 16;3(11):930–934.

28. Peisah C, et al. Pragmatic Systemic Solutions to the Wicked and Persistent Problem of the Unprofessional Disruptive Physician in the Health System. Healthcare (Basel). 2023 Sep 1;11(17):2455.

29. Vessey JA, et al. Bullying of staff registered nurses in the workplace: a preliminary study for developing personal and organizational strategies for the transformation of hostile to healthy workplace environments. J Prof Nurs. 2009 Sep-Oct;25(5):299–306.

30. Villafranca A, et al. Disruptive behaviour in the perioperative setting: a contemporary review. Can J Anaesth. 2017 Feb;64(2):128–140.

31. Jönsson S, Muhonen T. Factors influencing the behavior of bystanders to workplace bullying in healthcare-A qualitative descriptive interview study. Res Nurs Health. 2022 Aug;45(4):424–432.

32. Lang M, et al. Workplace bullying, burnout and resilience amongst perioperative nurses in Australia: A descriptive correlational study. J Nurs Manag. 2022 Sep;30(6):1502–1513.

33. Johnson AH, Benham-Hutchins M. The Influence of Bullying on Nursing Practice Errors: A Systematic Review. AORN J. 2020 Feb;111(2):199–210.

34. Hawkins N, Jeong S, Smith T. New graduate registered nurses’ exposure to negative workplace behaviour in the acute care setting: An integrative review. Int J Nurs Stud. 2019 May;93:41–54.

35. Edmonson C, Zelonka C. Our Own Worst Enemies: The Nurse Bullying Epidemic. Nurs Adm Q. 2019 Jul-Sep;43(3):274–279.

36. Boniol M, et al. The global health workforce stock and distribution in 2020 and 2030: a threat to equity and ‘universal’ health coverage?. BMJ Glob Health. 2022 Jun;7(6):e009316. doi: 10.1136/bmjgh-2022-009316.

37. Beal DJ, et al. Cohesion and performance in groups: a meta-analytic clarification of construct relations. J Appl Psychol. 2003 Dec;88(6):989–1004.

38. Mulder LB, et al. Undermining trust and cooperation: The paradox of sanctioning systems in social dilemmas. 2006 March;42(2):147–162.

39. Mazzetti G, Schaufeli WB. The impact of engaging leadership on employee engagement and team effectiveness: A longitudinal, multi-level study on the mediating role of personal- and team resources. PLOS One. 2022 Jun 29;17(6):e0269433. doi: 10.1371/journal.pone.0269433.

40. Zehr H, Lenses C. A. New focus for crime and justice, Virginia: Herald Press, 1990.

